# Risk of tuberculosis and uptake rates of latent tuberculosis infection screening among clinical risk groups in South Korea: A nationwide population-based cohort study

**DOI:** 10.1101/2023.06.02.23290863

**Authors:** Hyung Woo Kim, Jinsoo Min, Joon Young Choi, Ah Young Shin, Jun-Pyo Myong, Yunhee Lee, Hyeon Woo Yim, Hyunsuk Jeong, Sanghyuk Bae, Choi Hoyong, Hyekyung In, Ahyoung Park, Miri Jang, Hyeon-Kyoung Koo, Sung-Soon Lee, Jae Seuk Park, Ju Sang Kim

## Abstract

**Background:** This study aimed to investigate actual tuberculosis (TB) risk and uptake rates of latent tuberculosis infection (LTBI) screening among eight clinical risk groups specified in Korean guidelines. Proportions of potentially preventable TB in these groups were also calculated.

**Methods and Findings:** Patients enrolled before January 1^st^, 2018, were classified into a prevalence cohort whereas those enrolled thereafter were classified into an incidence cohort. Both cohorts were followed up until December 31^st^, 2020. Sex, age, and calendar year-adjusted standardized incidence ratio (SIR) of tuberculosis was calculated with total population in South Korea as a reference group. The number of TB patients notified in 2018 was investigated for both prevalence and incidence cohorts. SIR of TB in each incidence cohort was higher than that in each corresponding prevalence cohort. Among all incidence cohorts, SIR in people living with human immunodeficiency virus (PLHIV) was the highest (17.41, (95% CI: 14.14-21.43)). Although classified as moderate TB risk diseases in current guideline, end-stage renal disease (ESRD) (8.05, (7.02-9.23)) and uncontrolled diabetes mellitus (DM) (6.31, (5.78-6.99)) showed high SIRs comparable to other high-risk diseases. Among total TB cases notified in 2018, each cohort accounted for less than 1.5% except for patients with DM. The uptake rate of LTBI test was the highest among patients using TNF inhibitors (92.7%), followed by those who underwent organ transplantation (60.4%) and PLHIV (41.3%).

**Conclusions:** LTBI screening should be reinforced for certain clinical risk groups such as ESRD or uncontrolled DM. Beyond the current guideline, additional high-risk groups should be identified.

## INTRODUCTION

Approximately a quarter of population in the world is presumed to be infected with tuberculosis (TB)[1]. For TB elimination, strategies to tackle the large TB reservoir are essential[2]. However, current diagnostic tools for latent tuberculosis infection (LTBI) have low predictive values[3], which can result in high numbers needed treat to prevent a TB case. Several conditions raising TB risk have been investigated[4]. Previous guidelines on LTBI have commonly specified two key groups: TB contacts and clinical risk groups[5-8].

TB incidence in South Korea has continuously decreased since 2000. TB incidence was 96.3 cases per 100,000 population in 2000 and 44.6 cases per 100,000 population in 2021[9]. With a decrease in TB incidence, strategy to prevent reactivation has been underscored as in other low-incidence countries[10,11]. However, in contrast to contact investigation which is managed by the government[12], LTBI screening in clinical risk group is usually performed in private sector, which accounts for more than 90% healthcare facilities in South Korea[13]. Although clinical risk groups have been specified in Korean guidelines since amendment in 2014[14], actual TB risk and uptake rate of LTBI screening in these groups have not been evaluated before.

Thus, the objective of this study was to investigate the actual TB risk among high-risk groups specified in Korean guidelines for TB and uptake rate of LTBI screening in each group. Additionally, proportions of potentially preventable TB by implementing LTBI screening in these risk groups were calculated among total nationwide TB burden.

## METHODS

### Study population and data source

Eight diseases specified in Korean guidelines were selected. Records of patients with such disease were extracted from National Health Information Database (NHID) according to the operational definition of each disease (S1 Table). Each group was denoted as follows: Group 1, people living with human immunodeficiency virus (PLHIV); Group 2, patients who underwent solid organ or hematopoietic stem cell transplantation; Group 3, patients who used tumor necrosis factor (TNF) inhibitors; Group 4, patients with end-stage renal disease (ESRD); Group 5, patients who underwent gastrectomy; Group 6, patients with head & neck cancer; Group 7, patients with hematologic malignancy; and Group 8, patients with diabetes mellitus (DM). Additionally, in Group 8, patients who needed regular insulin injection implying patients with poorly controlled DM were selected and analyzed as Group 8-1. Three diseases (Group 1, Group 2, and Group 3) were classified as high-risk diseases while others were classified as moderate-risk diseases. The date of enrollment was defined as the first date that criteria for operational definition were all met. Patients enrolled before January 1^st^, 2018 were classified into a prevalence cohort, representing patients who had the disease at the timepoint of January 1^st^, 2018. Those enrolled on 1^st^ January 2018 or thereafter were classified into an incidence cohort, who were newly diagnosed patients. Patients died before January 1^st^, 2018 were excluded. Patients with missing information on demographic variables such as age and sex were also excluded. Each cohort was linked to database of Korean National TB Surveillance System (KNTSS) using national identification number.

### Study design

Patients enrolled in each cohort were followed up from January 1^st^, 2018 for the prevalence cohort and from the date of enrollment for the incidence cohort. Follow-up was terminated at 1) date of TB diagnosis, 2) date of death or 3) 31st December 2020, whichever came first. Incidence rate of TB and mortality were calculated for each cohort. To estimate TB risk, sex, age, and calendar year-adjusted standardized incidence ratio (SIR) of tuberculosis were calculated with total population in South Korea as a reference group. TB incidence among total population was calculated using database of KNTSS notified from 2018 to 2020. SIR was presented after stratifying subjects into three age groups -patients aged under 35 years, those aged between 35-64 years and those aged 65 years or over.

Age-stratified proportion of each prevalence and incidence cohort among nationwide TB patients notified in 2018 was calculated. For the incidence cohort, only patients enrolled in 2018 were analyzed.

The percentage of those who underwent interferon-gamma release assay (IGRA) or tuberculin skin test (TST) was investigated. Screening was classified by the date of LTBI examination. Recent screening denoted that the screening was performed within one year before the date of enrollment. New screening indicated that the screening was performed at the date of enrollment or thereafter. Past screening represented that the screening was performed more than one year before the date of enrollment. As some patients such as those who were scheduled for organ transplantation and those who were going to use TNF inhibitors were requested to undergo LTBI screening in advance, the percentage of recent or new screening was presented as one of the major outcomes of this analysis.

### Statistical analysis

SIR based on Poisson model with 95% confidence interval from Wald’s normal approximation was calculated using R package ‘popEpi’. All statistical analyses were conducted with R v.3.6.2 (R foundation for Statistical Computing, Vienna, Austria) and SAS software version 9.4 (SAS Institute Inc., Cary, NC, USA). Statistical significance was considered when two-sided *P-*value was less than 0.05.

### Ethics statement

The present study protocol was reviewed and approved by the Institutional Review Board (IRB) of Incheon St. Mary’s Hospital, the Catholic University of Korea (IRB No. OC20ZNDE0023). Korea Disease Control and Prevention Agency collected informed consent from all notified TB patients when they were enrolled according to Tuberculosis Prevention Act. Informed consent from enrolled patients with high or moderate TB risk disease was waived due to the retrospective nature of this study. All enrolled patients were anonymized.

## RESULTS

Numbers of finally included patients in prevalence and incidence cohorts are presented in Figure 1. Demographic features of enrolled patients in each group were demonstrated in S2 Table. Among eight prevalence cohorts, Group 4 showed the highest TB incidence (487.4 per 100,000 person-years) and mortality rate (9592.9 per 100,000 person-year). Similarly, among incidence cohorts, TB incidence (1117.4 per 100,000 person-years) and mortality (19619.5 per 100,000 person-years) were the highest in Group 4 (Table 1). In every group, TB incidence was higher in the incidence cohort than in the prevalence cohort. Mortality was higher in the incidence cohort than in the prevalence cohort for all groups except for Group 8.

**Fig 1.**
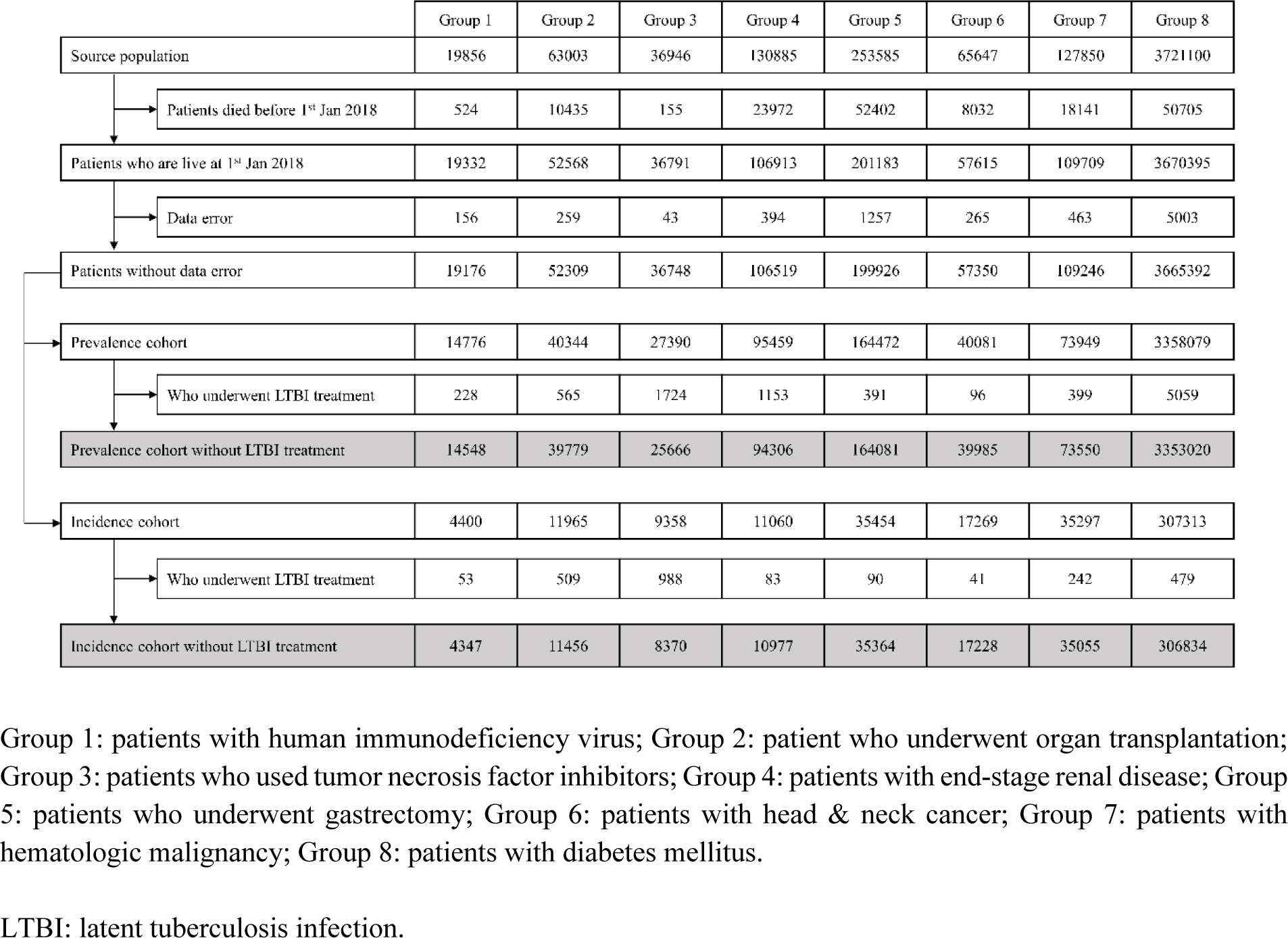
Flow diagram.

**Table 1.** Incidence of TB and death during follow-up period (2018-2020) among each prevalence cohort and each incidence cohort

|  | Prevalence cohort |  |  |  | Incidence cohort |  |  |  |
| --- | --- | --- | --- | --- | --- | --- | --- | --- |
|  | TB, N<br>(Incidence per<br>100000 person-years) | Death, N<br>(Incidence per<br>100000 person-years) | Total N | Total follow-up (person-years) | TB, N<br>(Incidence per<br>100000 person-years) | Death, N<br>(Incidence per<br>100000 person-years) | Total N | Total<br>follow-up<br>(person-years) |
| Group 1 | 106 (247.1) | 360 (839.2) | 14548 | 42898.8 | 89 (1068.3) | 162 (1944.6) | 4347 | 8330.8 |
| Group 2 | 167 (146.1) | 2794 (2444.6) | 39779 | 114293.7 | 105 (518.1) | 1867 (9211.7) | 11456 | 20267.6 |
| Group 3 | 135 (177.2) | 385 (505.3) | 25666 | 76188.8 | 75 (448.9) | 88 (526.7) | 8370 | 16706.2 |
| Group 4 | 1189 (487.4) | 23401 (9592.9) | 94306 | 243940.4 | 206 (1117.4) | 3617 (19619.5) | 10977 | 18435.7 |
| Group 5 | 1161 (249.1) | 15396 (3303.1) | 164081 | 466104 | 225 (338.5) | 3900 (5867.3) | 35364 | 66470.2 |
| Group 6 | 241 (219.7) | 5860 (5341.1) | 39985 | 109714.8 | 172 (575.5) | 3239 (10837.4) | 17228 | 29887.2 |
| Group 7 | 311 (153.1) | 9496 (4674.9) | 73550 | 203127.5 | 343 (620.5) | 9341 (16897.0) | 35055 | 55282.1 |
| Group 8 | 14677 (150.8) | 204247 (2098.1) | 3353020 | 9735057.0 | 1587 (208.8) | 12207 (1606.4) | 306834 | 759891.6 |
| Group 8-1 | 4881 (304.0) | 79825 (4971.9) | 580894 | 1605530.5 | 498 (571.8) | 5340 (6131.4) | 38136 | 87092.1 |

Group 1: patients with human immunodeficiency virus; Group 2: patient who underwent organ transplantation; Group 3: patients who used tumor necrosis factor inhibitors; Group 4: patients with end-stage renal disease; Group 5: patients who underwent gastrectomy; Group 6: patients with head & neck cancer; Group 7: patients with hematologic malignancy; Group 8: patients with diabetes mellitus; Group 8-1: patients with diabetes mellitus who needed regular insulin injection.
TB: tuberculosis.

### Standardized incidence ratio of TB

Age-stratified SIR of TB in the prevalence cohort is presented in Table 2. Group 4 had the highest SIR (3.76, 95% CI: 3.55-3.98), followed by Group 1 (3.61, 2.98-4.37), Group 3 (3.30, 2.79-3.90), and Group 8-1 (2.30, 2.24-2.37). Although there were subtle differences among cohorts, SIRs in elderly population were lower than those in younger population.

**Table 2.**
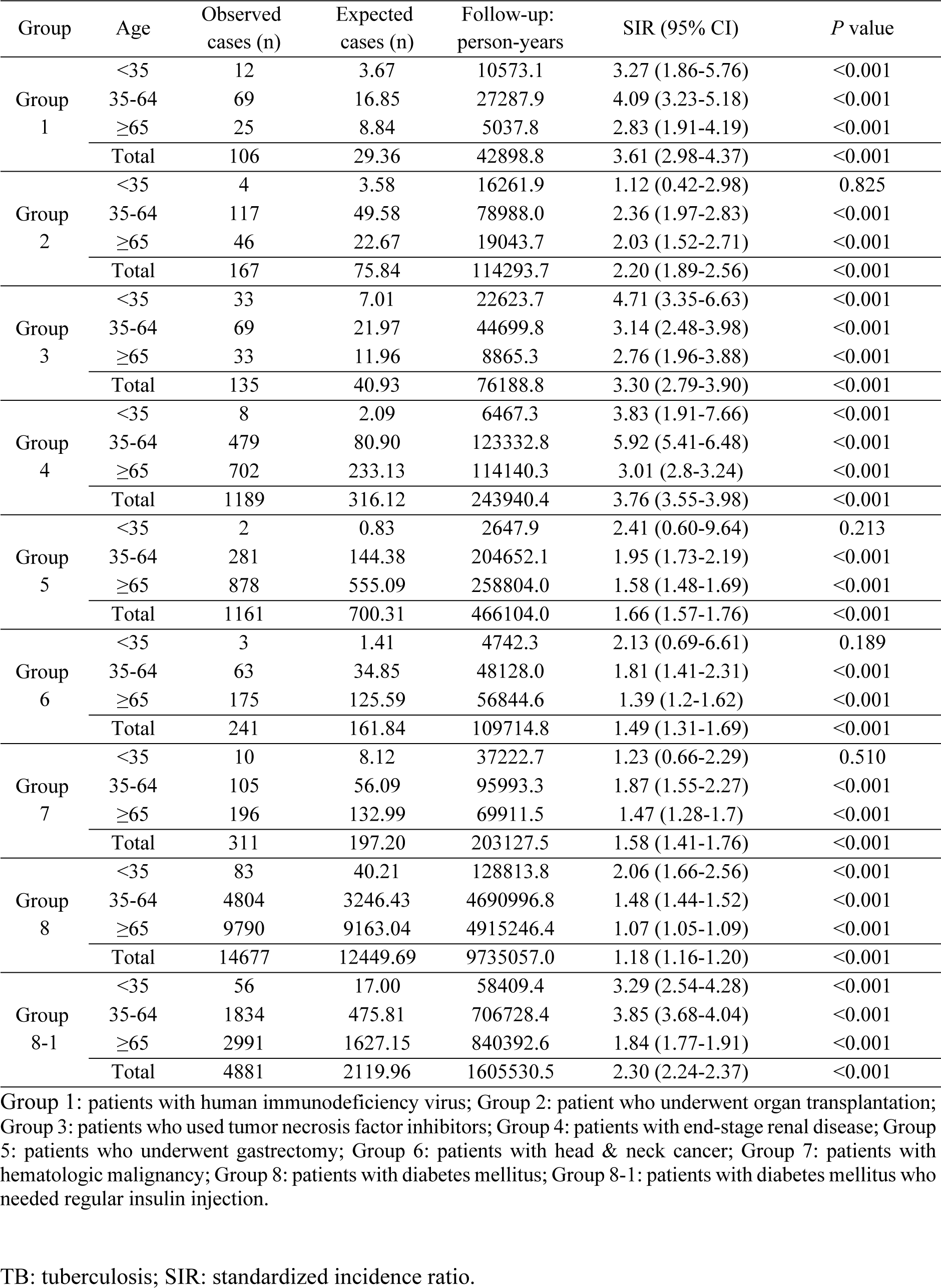
Standardized incidence ratio of TB among each prevalence cohort stratified by age

Among all groups, SIR of TB was higher in the incidence cohort than in the prevalence cohort (Table 3). Group 1 had the highest SIR (17.41, 14.14-21.43), followed by Group 3 (9.67, 7.71-12.12), Group 2 (8.90, 7.35-10.77), and Group 4 (8.05, 7.02-9.23), which comprised the high-TB risk group. Among other groups, Group 8-1 and Group 7 showed considerably high SIRs (6.31, 5.78-6.99 and 6.07, 5.46-6.75, respectively). As in the prevalence cohort, SIRs in elderly population were lower than those in younger population. This generation gap in SIR was prominent in Group 1. For incident Group 1, SIR was 33.43 (26.31-42.47) aged between 35 and 64 years and 1.42 (0.46-4.41) for those who aged 65 years or over. Group 7 also showed such generation gap. For incident Group 7, SIR was 22.27 (15.75-31.49) for those aged under 35 years and 4.65 (4.03-5.36) for those aged 65 years or over. However, in Group 3, such tendency was not prominent. For incident Group 3, SIRs for those aged under 35 years, aged between 35 and 64 years, and aged 65 years or over were 8.91 (5.37-14.78), 10.44 (7.69-14.18), and 8.84 (5.64-13.86), respectively.

**Table 3.** Standardized incidence ratio of TB among each incidence cohort stratified by age.

| Group | Age | Observed cases (n) | Expected cases (n) | Follow-up: person-years | SIR (95% CI) | P value |
| --- | --- | --- | --- | --- | --- | --- |
| Group 1 | <35 | 19 | 1.00 | 3192.6 | 19.03 (12.14-29.84) | <0.001 |
|  | 35-64 | 67 | 2.00 | 3972.8 | 33.43 (26.31-42.47) | <0.001 |
|  | ≥65 | 3 | 2.11 | 1165.4 | 1.42 (0.46-4.41) | 0.542 |
|  | Total | 89 | 5.11 | 8330.8 | 17.41 (14.14-21.43) | <0.001 |
| Group 2 | <35 | 9 | 0.59 | 3109.4 | 15.16 (7.89-29.13) | <0.001 |
|  | 35-64 | 77 | 8.75 | 14784.4 | 8.80 (7.04-11.00) | <0.001 |
|  | ≥65 | 19 | 2.46 | 2373.8 | 7.73 (4.93-12.12) | <0.001 |
|  | Total | 105 | 11.80 | 20267.6 | 8.90 (7.35-10.77) | <0.001 |
| Group 3 | <35 | 15 | 1.68 | 6557.5 | 8.91 (5.37-14.78) | <0.001 |
|  | 35-64 | 41 | 3.93 | 8501.3 | 10.44 (7.69-14.18) | <0.001 |
|  | ≥65 | 19 | 2.15 | 1647.4 | 8.84 (5.64-13.86) | <0.001 |
|  | Total | 75 | 7.76 | 16706.2 | 9.67 (7.71-12.12) | <0.001 |
| Group 4 | <35 | 5 | 0.25 | 863.1 | 19.87 (8.27-47.73) | <0.001 |
|  | 35-64 | 78 | 5.54 | 8628.0 | 14.07 (11.27-17.57) | <0.001 |
|  | ≥65 | 123 | 19.80 | 8944.6 | 6.21 (5.21-7.41) | <0.001 |
|  | Total | 206 | 25.59 | 18435.7 | 8.05 (7.02-9.23) | <0.001 |
| Group 5 | <35 | 0 | 0.20 | 675.0 | 0 |  |
|  | 35-64 | 86 | 21.84 | 33185.8 | 3.94 (3.19-4.86) | <0.001 |
|  | ≥65 | 139 | 60.71 | 32609.4 | 2.29 (1.94-2.70) | <0.001 |
|  | Total | 225 | 82.75 | 66470.2 | 2.72 (2.39-3.10) | <0.001 |
| Group 6 | <35 | 2 | 0.43 | 1705.0 | 4.62 (1.16-18.48) | 0.030 |
|  | 35-64 | 75 | 9.46 | 14202.3 | 7.92 (6.32-9.94) | <0.001 |
|  | ≥65 | 95 | 28.74 | 13979.9 | 3.31 (2.70-4.04) | <0.001 |
|  | Total | 172 | 38.63 | 29887.2 | 4.45 (3.83-5.17) | <0.001 |
| Group 7 | <35 | 32 | 1.44 | 7470.2 | 22.27 (15.75-31.49) | <0.001 |
|  | 35-64 | 122 | 14.40 | 26056.1 | 8.47 (7.09-10.12) | <0.001 |
|  | ≥65 | 189 | 40.66 | 21755.7 | 4.65 (4.03-5.36) | <0.001 |
|  | Total | 343 | 56.50 | 55282.1 | 6.07 (5.46-6.75) | <0.001 |
| Group 8 | <35 | 38 | 10.95 | 36272.6 | 3.47 (2.53-4.77) | <0.001 |
|  | 35-64 | 912 | 308.16 | 504621.9 | 2.96 (2.77-3.16) | <0.001 |
|  | ≥65 | 637 | 367.35 | 218997.1 | 1.73 (1.60-1.87) | <0.001 |
|  | Total | 1587 | 686.46 | 759891.6 | 2.31 (2.20-2.43) | <0.001 |
| Group 8-1 | <35 | 15 | 2.84 | 10598.4 | 5.29 (3.19-8.77) | <0.001 |
|  | 35-64 | 308 | 31.59 | 53195.6 | 9.75 (8.72-10.9) | <0.001 |
|  | ≥65 | 175 | 44.56 | 23298.2 | 3.93 (3.39-4.55) | <0.001 |
|  | Total | 498 | 78.98 | 87092.1 | 6.31 (5.78-6.88) | <0.001 |

Group 1: patients with human immunodeficiency virus; Group 2: patient who underwent organ transplantation; Group 3: patients who used tumor necrosis factor inhibitors; Group 4: patients with end-stage renal disease; Group 5: patients who underwent gastrectomy; Group 6: patients with head & neck cancer; Group 7: patients with hematologic malignancy; Group 8: patients with diabetes mellitus; Group 8-1: patients with diabetes mellitus who needed regular insulin injection.
TB: tuberculosis; SIR: standardized incidence ratio.

### Proportion of each cohort among nationwide TB cases

Table 4 presents number of TB cases notified in 2018 among each cohort and proportion of each cohort among 33,328 nationwide TB cases notified in 2018. Most of prevalence cohorts accounted for less than 1.5% of total notified TB cases except for Group 8 (17.2%). The proportion of each incidence cohort was similar to or lower than that of the prevalence cohort. Most of incidence cohorts accounted for less than 1% of total notified TB cases except for Group 8 (3.3%).

**Table 4.** Number of TB cases notified in South Korea, 2018 among each cohort and proportion of each cohort among total nationwide TB cases notified in South Korea, 2018 (n = 33,328)

|  | TB cases among prevalence cohort,<br>N (proportion %) | TB cases among incidence cohort<sup>a</sup>,<br>N (proportion %) |
| --- | --- | --- |
| **All age (n=33,328)** |  |  |
| Group 1 | 48 (0.1) | 50 (0.2) |
| Group 2 | 80 (0.2) | 34 (0.1) |
| Group 3 | 79 (0.2) | 26 (0.1) |
| Group 4 | 501 (1.5) | 88 (0.3) |
| Group 5 | 480 (1.4) | 89 (0.3) |
| Group 6 | 103 (0.3) | 64 (0.2) |
| Group 7 | 144 (0.4) | 158 (0.5) |
| Group 8 | 5724 (17.2) | 1097 (3.3) |
| Group 8-1 | 2453 (7.4) | 398 (1.2) |
| Total | 6613 (19.8) | 1572 (4.7) |
| **Age <35 (n=4,155)** |  |  |
| Group 1 | 6 (0.1) | 8 (0.2) |
| Group 2 | 3 (0.1) | 4 (0.1) |
| Group 3 | 21 (0.5) | 7 (0.2) |
| Group 4 | 6 (0.1) | 2 (0.0) |
| Group 5 | 3 (0.1) | 0 (0) |
| Group 6 | 2 (0.0) | 1 (0.0) |
| Group 7 | 10 (0.2) | 18 (0.4) |
| Group 8 | 38 (0.9) | 24 (0.6) |
| Group 8-1 | 29 (0.7) | 12 (0.3) |
| Total | 84 (2.0) | 58 (1.4) |
| **Age 35-64 (n=13,916)** |  |  |
| Group 1 | 35 (0.3) | 38 (0.3) |
| Group 2 | 61 (0.4) | 26 (0.2) |
| Group 3 | 39 (0.3) | 14 (0.1) |
| Group 4 | 203 (1.5) | 37 (0.3) |
| Group 5 | 140 (1.0) | 36 (0.3) |
| Group 6 | 37 (0.3) | 26 (0.2) |
| Group 7 | 55 (0.4) | 60 (0.4) |
| Group 8 | 2067 (14.9) | 689 (5.0) |
| Group 8-1 | 987 (7.1) | 258 (1.9) |
| Total | 2411 (17.3) | 913 (6.6) |
| **Age ≥ 65 (n=15,257)** |  |  |
| Group 1 | 7 (0.0) | 4 (0.0) |
| Group 2 | 16 (0.1) | 4 (0.0) |
| Group 3 | 19 (0.1) | 5 (0.0) |
| Group 4 | 292 (1.9) | 49 (0.3) |
| Group 5 | 337 (2.2) | 53 (0.3) |
| Group 6 | 64 (0.4) | 37 (0.2) |
| Group 7 | 79 (0.5) | 80 (0.5) |
| Group 8 | 3619 (23.7) | 384 (2.5) |
| Group 8-1 | 1437 (9.4) | 128 (0.8) |
| Total | 4118 (27.0) | 601 (3.9) |

<sup>a</sup>Only patients who were enrolled in 2018 were analyzed.
Group 1: patients with human immunodeficiency virus; Group 2: patient who underwent organ transplantation; Group 3: patients who used tumor necrosis factor inhibitors; Group 4: patients with end-stage renal disease; Group 5: patients who underwent gastrectomy; Group 6: patients with head & neck cancer; Group 7: patients with hematologic malignancy; Group 8: patients with diabetes mellitus; Group 8-1: patients with diabetes mellitus who needed regular insulin injection.
TB: tuberculosis.

Age-stratified proportion of each cohort was presented in Table 4. For high TB-risk diseases (Group 1, 2, 3) and DM (Group 8) proportions of incidence cohort were highest among TB patients aged between 35-64 years. For moderate TB-risk diseases such as ESRD (Group 4), or malignancy (Group 5, 6, 7), those were highest among TB patients aged over 65 years. Proportions of each incidence cohort were relatively low among TB patients aged under 35 years, in most of diseases. However, those of high TB-risk disease (Group 1, 2, 3) among TB patient aged over 65 years were even lower than among TB patients aged under 35 years.

### LTBI test uptake rate

The uptake rate of LTBI test was the highest in Group 3, with 92.7% of patients in Group 3 undergoing LTBI test within a year before or after the initiation of TNF inhibitors (Table 5). The percentage was 41.3% in Group 1 and 60.4% in Group 2. These percentages were lower in groups with moderate TB risk diseases. Only 14.1% and 12.0% of patients in Group 4 and Group 7 underwent LTBI test recently or newly, respectively. These percentage for other moderate TB risk groups were even lower. IGRA rather than TST was the mostly used LTBI test in each group.

**Table 5.** Percentage of patients who underwent latent TB screening with interferon-gamma release assay (IGRA) or tuberculin skin test (TST) for each incidence cohort.

| (1) IGRA or TST |  |  |  |  |
| --- | --- | --- | --- | --- |
|  | Recent or new screening (row %) | Past screening (row %) | No screening (row %) | Total |
| Group 1 | 1815 (41.3) | 43 (1.0) | 2542 (57.8) | 4400 |
| Group 2 | 7221 (60.4) | 315 (2.6) | 4429 (37.0) | 11965 |
| Group 3 | 8675 (92.7) | 358 (3.8) | 325 (3.5) | 9358 |
| Group 4 | 1558 (14.1) | 155 (1.4) | 9347 (84.5) | 11060 |
| Group 5 | 424 (1.2) | 223 (0.6) | 34807 (98.2) | 35454 |
| Group 6 | 393 (2.3) | 207 (1.2) | 16669 (96.5) | 17269 |
| Group 7 | 4235 (12.0) | 490 (1.4) | 30572 (86.6) | 35297 |
| Group 8 | 2103 (0.7) | 231 (0.1) | 304979 (99.2) | 307313 |
| Group 8-1 | 794 (2.1) | 53 (0.1) | 37386 (97.8) | 38233 |
| (2) IGRA |  |  |  |  |
|  | Recent or new IGRA (row %) | Past IGRA (row %) | No IGRA (row %) | Total |
| Group 1 | 1802 (41.0) | 24 (0.5) | 2574 (58.5) | 4400 |
| Group 2 | 7108 (59.4) | 265 (2.2) | 4592 (38.4) | 11965 |
| Group 3 | 8650 (92.4) | 335 (3.6) | 373 (4.0) | 9358 |
| Group 4 | 1551 (14.0) | 91 (0.8) | 9418 (85.2) | 11060 |
| Group 5 | 419 (1.2) | 127 (0.4) | 34908 (98.5) | 35454 |
| Group 6 | 390 (2.3) | 111 (0.6) | 16768 (97.1) | 17269 |
| Group 7 | 4193 (11.9) | 294 (0.8) | 30810 (87.3) | 35297 |
| Group 8 | 2067 (0.7) | 196 (0.1) | 305050 (99.3) | 307313 |
| Group 8-1 | 779 (2.0) | 43 (0.1) | 37411 (97.9) | 38233 |
| (3) TST |  |  |  |  |
|  | Recent or new TST (row %) | Past TST (row %) | No TST (row %) | Total |
| Group 1 | 56 (1.3) | 37 (0.8) | 4307 (97.9) | 4400 |
| Group 2 | 774 (6.5) | 264 (2.2) | 10927 (91.3) | 11965 |
| Group 3 | 919 (9.8) | 478 (5.1) | 7961 (85.1) | 9358 |
| Group 4 | 84 (0.8) | 98 (0.9) | 10878 (98.4) | 11060 |
| Group 5 | 15 (0.0) | 112 (0.3) | 35327 (99.6) | 35454 |
| Group 6 | 7 (0.0) | 119 (0.7) | 17143 (99.3) | 17269 |
| Group 7 | 127 (0.4) | 270 (0.8) | 34900 (98.9) | 35297 |
| Group 8 | 126 (0.0) | 72 (0.0) | 307115 (99.9) | 307313 |
| Group 8-1 | 63 (0.2) | 23 (0.1) | 38147 (99.8) | 38233 |

Group 1: patients with human immunodeficiency virus; Group 2: patient who underwent organ transplantation; Group 3: patients who used tumor necrosis factor inhibitors; Group 4: patients with end-stage renal disease; Group 5: patients who underwent gastrectomy; Group 6: patients with head & neck cancer; Group 7: patients with hematologic malignancy; Group 8: patients with diabetes mellitus; Group 8-1: patients with diabetes mellitus who needed regular insulin injection.
IGRA: interferon-gamma release assay; TST: tuberculin skin test.

Recent screening denotes that latent TB examination (IGRA or TST) is performed within one year before the date of enrollment. New screening indicates that the screening is performed at the date of enrollment or thereafter. Past screening represents that the screening is performed more than one year before the date of enrollment.

## DISCUSSION

In this study, well-known high TB risk diseases showed high SIRs as expected. Patients with ESRD showed TB risk comparable to those with high TB risk diseases. TB risk was relatively low in patients who underwent gastrectomy and patients with DM among groups with moderate TB risk. However, patients with uncontrolled DM status showed a relatively high TB risk. The uptake rate of LTBI screening in these clinical risk group was still suboptimal in all groups except for patients who used TNF inhibitors.

Previous LTBI guidelines did not specify when to treat in detail[5-8,15]. We demonstrated that TB risk of patients who developed the disease newly was higher than that of those who were diagnosed several years ago. This might be attributable to a decreased immunity of patients with an uncontrolled status of disease around the date of diagnosis. Considering that in South Korea, diagnosis rate of human immunodeficiency virus (HIV) was still suboptimal[16]. Many patients presented low CD4+ cell levels at diagnosis[17], which might have increased their vulnerability to TB at diagnosis as shown in a previous study[18]. Uncontrolled naïve DM and uremic status of patients with chronic kidney disease (CKD) initiating dialysis might have also contributed to the high TB incidence at diagnosis. Additionally, intensive immunosuppressive treatment following solid organ or hematopoietic stem cell transplantation and consecutive use of TNF inhibitors could increase TB risk. Among patients with malignancy, it was presumed that immunosuppressive states might be temporary during a session of anticancer treatments such as chemotherapy just after the diagnosis of cancer, which was demonstrated by decreasing SIR of TB with increasing time after cancer diagnosis[19].

Our findings underscored that LTBI screening should be focused on incident groups. However, most of incident groups showed higher mortality as well as higher TB incidence than prevalent groups. Among patients who were newly diagnosed as ESRD, 33.0% (3617/10977) of patients died during the follow-up period. Considering adverse effects of LTBI treatment such as gastrointestinal trouble and hepatotoxicity[20], we speculate that providing LTBI treatment to these critically ill patients might be unfeasible in many cases. Moreover, in patients newly diagnosed as malignancy, preventing TB would not be a medical priority. In a previous study, TB incidence was higher among patients with malignancy such as esophageal, lung, pancreatic cancer, and multiple myeloma, showing a relatively low 5-year survival[21]. We expect that the acceptance rate for LTBI treatment among these patients with limited life expectancy would be low. Low LTBI screening uptake rate in patients with malignancy (Groups 5, 6, 7) might be related to this reason.

However, Groups 2 and 3 showed relatively high uptake rates for LTBI screening (60.4% and 92.7%, respectively). This might be because LTBI screening and treatment are more feasible in these groups than in other groups. Mortality rate of Group 3 was the lowest among all incident groups. This finding suggests that LTBI screening would be more feasible when they are scheduled to be a high-risk group, than when they had already become. Relatively low uptake rate of LTBI screening rate among PLHIV (41.3%) who were specified as the highest TB risk group could be explained by this hypothesis considering a low diagnosis rate of HIV and a low CD4+ cell count at diagnosis suggesting delayed diagnosis of HIV in South Korea[16,17]. We presume that diagnosis of HIV in earlier course of the disease would enhance the LTBI screening uptake rate. Further studies are needed to verify this hypothesis.

ESRD showed high SIR as other diseases with high TB risk, although it was specified as a disease with moderate TB risk in South Korea[15]. Additionally, it showed the highest SIR among all prevalent groups, suggesting that immunosuppression state could last for a longer period than other groups. Therefore, it should be reclassified as a disease with a high TB risk. However, LTBI screening is not widely performed for those with ESRD. We speculate that screening LTBI at earlier stage in chronic kidney disease (CKD) could be an alternative option for increasing the uptake rate of LTBI screening, like that for PLHIV. However, potential nephrotoxicity of anti-TB medication, especially rifampicin, is a concern[22], which might lower the uptake rate of LTBI treatment among patients with pre-dialysis CKD. LTBI regimen without potential risk of nephrotoxicity should be investigated.

DM is a known risk factor for TB[23]. However, in our study, TB risk was relatively low among diseases with moderate TB risk. Instead, uncontrolled DM status rather than disease itself contributed to TB development, as reported in a previous study[24]. By focusing on patients with uncontrolled DM, the cost of LTBI screening could be reduced while the effectiveness of TB prevention is increased. However, mortality rate of this group was quite high – more than 10 times as high as that of the total DM group. This suggests that there might be other serious comorbidities among them. As in patients with ESRD and those with malignancy, comorbidities might limit the feasibility of LTBI screening.

Besides feasibility, strategy targeting these clinical risk groups has other limitations. First, these clinical risk groups accounted for only small proportion of nationwide TB patients – less than 1.5% for each group except for patients with DM. Similarly to our results, Ronald et al. have demonstrated that application of WHO’s LTBI guideline targeting only for TB contacts and clinical risk group can only minimally impact the national TB incidence of Canada[25]. Only 4.5% of active TB cases were preventable with this strategy. This suggests that covering clinical risk group is essential but insufficient to reduce TB burden. Identification of additional high-risk groups is required[26]. Second, when compared to SIR in younger generation, SIR in elderly population was much lower in most groups. This suggests that identifying diseases with a high TB risk could be a useful tool for risk stratification among younger generation whereas it is less useful in elderly population. The low SIR in elderly population is attributable to a high TB incidence among general elderly population in South Korea[27]. Other comorbidities not specified in current guidelines, waning immunity derived from aging, and malnutrition might also contribute to this[28]. In addition, we demonstrated that proportions of high TB risk diseases among elderly TB patients were extremely low, and lower than other age groups, implying LTBI screening strategy targeting for high TB risk diseases is less efficient in elderly population, than in other age groups.

In the previous Canadian study, covering immigrants from high TB burden countries potentially prevented 37.1% of total TB cases[25]. However, in South Korea, the proportion of foreign-born residents among total population was 3.4% in 2020, which was lower than average (14.7%) of other high-income countries[29]. Instead, native elderly TB patients are a key group in South Korea. In 2021, TB patients who aged 65 years or above accounted for approximately 51.0% of total TB cases[9]. LTBI screening and treatment among elderly population are not routinely recommended due to low predictive values of diagnostic tools such as IGRA and TST and higher incidence of adverse effects during LTBI treatment[20]. Recently, the necessity of expanding LTBI screening to elderly population has been suggested[30] and the feasibility of LTBI treatment among high-risk elderly population has been reported[31]. Further studies identifying high-risk elderly population should be implemented.

This study has a strength of linking two databases covering the entire South Korean population. Thus, many study subjects were included. We compared several diseases with high or moderate TB risk simultaneously at nationwide level within the same study period. However, our study had a limitation in that the LTBI status of each patient was unavailable. SIR among patients with LTBI might reflect more accurate TB risk. However, considering that most diseases did not affect LTBI prevalence[5] and that age was the most significant factor associated with LTBI prevalence[32], we assumed that LTBI prevalence in each group and general population were not quite different, and that age-adjusted SIR would be sufficient for estimating the actual TB risk. Another limitation was that the number of patients with LTBI was not known. Thus, calculation of further cascade of care such as initiation of LTBI treatment was unfeasible.

In conclusion, LTBI screening in certain clinical risk groups such as patients with ESRD and patients with uncontrolled DM should be reinforced. Ideally, LTBI screening should be provided around the date of the disease diagnosis. However, feasibility of LTBI treatment at that period remains a problem. Beyond the current guideline, identification of additional high-risk groups, especially among elderly population, is required.

## Data Availability

Data cannot be shared publicly, because personal information of Korean population in included in these datasets, which are protected under Personal Information Protection Act of South Korea. Korea Disease Control and Prevention Agency (KDCA) and National Health Insurance Service of Korea (NHIS) owns all datasets. The data used in the current study are available only after the permission from the KDCA and NHIS in advance.

## References

1. Houben RMGJ, Dodd PJ. The Global Burden of Latent Tuberculosis Infection: A Re-estimation Using Mathematical Modelling. PLOS Medicine. 2016; 13:e1002152. https://doi.org/10.1371/journal.pmed.1002152

2. Dye C, Glaziou P, Floyd K, Raviglione M. Prospects for tuberculosis elimination. Annu Rev Public Health. 2013; 34:271–286. https://doi.org/10.1146/annurev-publhealth-031912-114431 PMID: 23244049

3. Rangaka MX, Wilkinson KA, Glynn JR, Ling D, Menzies D, Mwansa-Kambafwile J, et al. Predictive value of interferon-γ release assays for incident active tuberculosis: a systematic review and meta-analysis. Lancet Infect Dis. 2012; 12:45–55. https://doi.org/10.1016/s1473-3099(11)70210-9 PMID: 21846592

4. Leung CC, Rieder HL, Lange C, Yew WW. Treatment of latent infection with Mycobacterium tuberculosis: update 2010. Eur Respir J. 2011; 37:690–711. https://doi.org/10.1183/09031936.00079310 PMID: 20693257

5. European Centre for Disease Prevention and Control. Programmatic management of latent tuberculosis infection in the European Union; 2018 [cited 2023 25th Jan]. Available from: https://www.ecdc.europa.eu/en/publications-data/programmatic-management-latent-tuberculosis-infection-european-union

6. National Institute for Health and Care Excellence. Tuberculosis, NICE guideline; 2016 [cited 2023 25th Jan]. Available from: www.nice.org.uk/guidance/ng33

7. World Health Organization. WHO consolidated guidelines on tuberculosis: module 1: prevention: tuberculosis preventive treatment; 2020 [cited 2023 25th Jan]. Available from: https://www.who.int/publications/i/item/9789240001503

8. Targeted tuberculin testing and treatment of latent tuberculosis infection. Am J Respir Crit Care Med. 2000; 161:S221-247. https://doi.org/10.1164/ajrccm.161.supplement_3.ats600 PMID: 10764341

9. Korea Disease Control and Prevention Agency. Annual Report on the Notified Tuberculosis in Korea, 2021. 2022.

10. Shea KM, Kammerer JS, Winston CA, Navin TR, Horsburgh CR, Jr. Estimated rate of reactivation of latent tuberculosis infection in the United States, overall and by population subgroup. Am J Epidemiol. 2014; 179:216–225. https://doi.org/10.1093/aje/kwt246 PMID: 24142915

11. World Health Organization. Towards tuberculosis elimination: an action framework in low-incidence countries. Geneva; 2014.

12. Go U, Park M, Kim U-N, Lee S, Han S, Lee J, et al. Tuberculosis prevention and care in Korea: Evolution of policy and practice. Journal of Clinical Tuberculosis and Other Mycobacterial Diseases. 2018; 11:28–36. https://doi.org/10.1016/j.jctube.2018.04.006

13. Organization for Economic Cooperation and Development. OECD Reviews of Public Health: Korea. A Healthier Tomorrow; 2020. Available from: https://www.oecd-ilibrary.org/sites/6e005d47-en/index.html?itemId=/content/component/6e005d47-en

14. Joint Committee for the Revision of Korean Guidelines for Tuberculosis. Korean Guidelines for Tuberculosis, 2nd edition. Korea Centers for Disease Control and Prevention; 2014.

15. Joint Committee for the Revision of Korean Guidelines for Tuberculosis. Korean Guidelines for Tuberculosis, 4th edition. Korea Centers for Disease Control and Prevention; 2020.

16. Lee E, Kim J, Lee JY, Bang JH. Estimation of the Number of HIV Infections and Time to Diagnosis in the Korea. J Korean Med Sci. 2020; 35:e41. https://doi.org/10.3346/jkms.2020.35.e41 PMID: 32056401

17. Kim MJ, Chang HH, Kim SI, Kim YJ, Park DW, Kang C, et al. Trend of CD4+ Cell Counts at Diagnosis and Initiation of Highly Active Antiretroviral Therapy (HAART): Korea HIV/AIDS Cohort Study, 1992-2015. Infect Chemother. 2017; 49:101-108. https://doi.org/10.3947/ic.2017.49.2.101 PMID: 28608664

18. Ellis PK, Martin WJ, Dodd PJ. CD4 count and tuberculosis risk in HIV-positive adults not on ART: a systematic review and meta-analysis. PeerJ. 2017; 5:e4165. https://doi.org/10.7717/peerj.4165 PMID: 29259846

19. Cheng MP, Abou Chakra CN, Yansouni CP, Cnossen S, Shrier I, Menzies D, et al. Risk of Active Tuberculosis in Patients with Cancer: A Systematic Review and Meta-Analysis. Clin Infect Dis. 2017; 64:635–644. https://doi.org/10.1093/cid/ciw838 PMID: 27986665

20. Campbell JR, Dowdy D, Schwartzman K. Treatment of latent infection to achieve tuberculosis elimination in low-incidence countries. PLoS Med. 2019; 16:e1002824. https://doi.org/10.1371/journal.pmed.1002824 PMID: 31170161

21. Cheon J, Kim C, Park EJ, Ock M, Lee H, Ahn JJ, et al. Active tuberculosis risk associated with malignancies: an 18-year retrospective cohort study in Korea. J Thorac Dis. 2020; 12:4950–4959. https://doi.org/10.21037/jtd.2020.02.50 PMID: 33145069

22. Grilo Novais A, Silva C, Coelho AR, Silva R, Carvalho AC. Rifampicin-Induced Nephrotoxicity in a Tuberculosis Patient: Treatment Dilemma? Eur J Case Rep Intern Med. 2021; 8:002833. https://doi.org/10.12890/2021_002833 PMID: 34790626

23. Jeon CY, Murray MB. Diabetes mellitus increases the risk of active tuberculosis: a systematic review of 13 observational studies. PLoS Med. 2008; 5:e152. https://doi.org/10.1371/journal.pmed.0050152 PMID: 18630984

24. Yoo JE, Kim D, Han K, Rhee SY, Shin DW, Lee H. Diabetes Status and Association With Risk of Tuberculosis Among Korean Adults. JAMA Netw Open. 2021; 4:e2126099. https://doi.org/10.1001/jamanetworkopen.2021.26099 PMID: 34546370

25. Ronald LA, Campbell JR, Rose C, Balshaw R, Romanowski K, Roth DZ, et al. Estimated Impact of World Health Organization Latent Tuberculosis Screening Guidelines in a Region With a Low Tuberculosis Incidence: Retrospective Cohort Study. Clin Infect Dis. 2019; 69:2101–2108. https://doi.org/10.1093/cid/ciz188 PMID: 30856258

26. Bigio J, Viscardi A, Gore G, Matteelli A, Sulis G. A scoping review on the risk of tuberculosis in specific population groups: can we expand the World Health Organization recommendations? Eur Respir Rev. 2023; 32 https://doi.org/10.1183/16000617.0127-2022 PMID: 36631131

27. Kim JH, Yim JJ. Achievements in and Challenges of Tuberculosis Control in South Korea. Emerg Infect Dis. 2015; 21:1913–1920. https://doi.org/10.3201/eid2111.141894 PMID: 26485188

28. Caraux-Paz P, Diamantis S, de Wazières B, Gallien S. Tuberculosis in the Elderly. J Clin Med. 2021; 10 https://doi.org/10.3390/jcm10245888 PMID: 34945187

29. United Nations Department of Economic and Social Affairs Population Division. International Migrant Stock 2020. 2020.

30. Huynh GH, Klein DJ, Chin DP, Wagner BG, Eckhoff PA, Liu R, et al. Tuberculosis control strategies to reach the 2035 global targets in China: the role of changing demographics and reactivation disease. BMC Med. 2015; 13:88. https://doi.org/10.1186/s12916-015-0341-4 PMID: 25896465

31. Huang HL, Huang WC, Lin KD, Liu SS, Lee MR, Cheng MH, et al. Completion Rate and Safety of Programmatic Screening and Treatment for Latent Tuberculosis Infection in Elderly Patients With Poorly Controlled Diabetic Mellitus: A Prospective Multicenter Study. Clin Infect Dis. 2021; 73:e1252–e1260. https://doi.org/10.1093/cid/ciab209 PMID: 33677558

32. Kim HW, Min J, Choi JY, Shin AY, Myong JP, Lee Y, et al. Prevalence of latent tuberculosis infection among participants of the national LTBI screening program in South Korea - A problem of low coverage rate with current LTBI strategy. Front Public Health. 2022; 10:1066269. https://doi.org/10.3389/fpubh.2022.1066269 PMID: 36743163

